# Trajectories of Clinical and Laboratory Characteristics Associated with COVID-19 in Hemodialysis Patients by Survival

**DOI:** 10.1101/2021.02.28.21252383

**Authors:** Sheetal Chaudhuri, Rachel Lasky, Yue Jiao, John Larkin, Caitlin Monaghan, Anke Winter, Luca Neri, Peter Kotanko, Jeffrey Hymes, Sangho Lee, Yuedong Wang, Jeroen P. Kooman, Franklin Maddux, Len Usvyat

## Abstract

**Introduction:** The clinical impact of COVID-19 has not been established in the dialysis population. We evaluated the trajectories of clinical and laboratory parameters in hemodialysis (HD) patients.

**Methods:** We used data from adult HD patients treated at an integrated kidney disease company who received a RT-PCR test to investigate suspicion of a SARS-CoV-2 infection between 01 May and 01 Sep 2020. Nonparametric smoothing splines were used to fit data for individual trajectories and estimate the mean change over time in patients testing positive or negative for SARS-CoV-2 and those who survived or died within 30 days of first suspicion or positive test date. For each clinical parameter of interest, the difference in average daily changes between COVID-19 positive versus negative group and COVID-19 survivor versus non-survivor group was estimated by fitting a linear mixed effects model based on measurements in the 14 days before (i.e., day-14 to day 0) day 0.

**Results:** There were 12,836 HD patients with a suspicion of COVID-19 who received RT-PCR testing (8,895 SARS-CoV-2 positive). We observed significantly different trends (p<0.05) in pre-HD systolic blood pressure (SBP), pre-HD pulse rate, body temperature, ferritin, lymphocytes, albumin, and interdialytic weight gain (IDWG) between COVID-19 positive and negative patient. For COVID-19 positive group, we observed significantly different clinical trends (p<0.05) in pre-HD pulse rate, lymphocytes, albumin and neutrophil-lymphocyte ratio (NLR) between survivors and non-survivors. We also observed that, in the group of survivors, most clinical parameters returned to pre-COVID-19 levels within 60-90 days.

**Conclusion:** We observed unique temporal trends in various clinical and laboratory parameters among HD patients who tested positive versus negative for SARS-CoV-2 infection and those who survived the infection versus those who died. These trends can help to define the physiological disturbances that characterize the onset and course of COVID-19 in HD patients

## Introduction

The Coronavirus Disease (COVID-19) pandemic has greatly affected the dialysis community. Dialysis patients appear to be at increased risk for viral transmission with relatively high mortality rates ranging from 11% to 30% ^1-6^. During the first half of 2020, there were over 11,200 COVID-19 hospitalizations among Medicare beneficiaries undergoing dialysis in the United States ^7^. Various parameters such as pulse, body temperature, C-reactive protein (CRP) and lymphocyte counts at presentation were found to be associated to COVID-19 mortality in kidney failure ^4^. However, the incubation time has not been clearly defined and patients may be infected with Severe Acute Respiratory Syndrome Coronavirus-2 (SARS-CoV-2) and potentially infectious weeks before presentation with symptoms. Early detection of changes in physiological parameters have been suggested to aid the identification/prediction of patients at risk for COVID-19 ^8^. Apart from early detection, trends in clinical and laboratory parameters may also have prognostic significance. For example, distinct differences in trajectories of clinical and laboratory parameters before the start of kidney replacement therapy have been shown for patients who survived versus died during the first year on hemodialysis (HD) ^9^.

Albeit the clinical presentations in COVID-19 have been somewhat established ^4^, the changes in clinical parameters before presentation that characterize disease onset in humans are unknown secondary to a scarcity of longitudinal data available in the general population or collected in registries in the kidney failure population. HD patients have robust routine data collected in Electronic Health Records (EHRs) affording the opportunity to define the pathophysiological disturbances characterizing the onset and course of COVID-19 in kidney failure patients. The goal of this analysis was to compare trends in clinical and laboratory parameters between HD patients who tested positive or negative for SARS CoV-2. The second goal of this study was to compare clinical trends between survivors and non-survivors who were diagnosed with COVID-19.

## Methods

### General Design

We used data from HD patients treated at a dialysis network in the United States of an integrated kidney disease company (Fresenius Medical Care, Waltham, MA, United States) between May and November 2020. HD patients who were suspected to have a SARS-CoV-2 infection at the outpatient dialysis clinics universally received reverse transcription polymerase chain reaction (RT-PCR) testing to diagnosis COVID-19. For this analysis, we retrospectively evaluated the trends in clinical and laboratory parameters 90 days before the date of suspicion for SARS-CoV-2 infection among HD patients who were diagnosed RT-PCR COVID-19 positive versus those that were negative. Among HD patients with RT-PCR confirmed COVID-19, we also assessed at the trajectories for those who survived versus died within 30 days after suspicion of SARS-CoV-2 infection.

This analysis was performed under a protocol reviewed by New England Institutional Review Board (Needham Heights, MA, United States; Version 1.0 NEIRB# 17-1376378-1) who determined this analysis of existing patient data that was de-identified by the investigator was exempt and did not require informed consent. This analysis was conducted in adherence with the Declaration of Helsinki.

### Patient Population

We included data from adult (age ≥18 years) HD patients who received RT-PCR testing to investigate suspicion of a SARS-CoV-2 infection between 01 May and 01 September 2020. We required patients to have a minimum follow-up period of 90 days; follow-up data was captured through 30 November 2020 as applicable. Suspicion of SARS-CoV-2 infection was determined at presentation by active signs and symptoms of a flu-like illness. We excluded data from patients under investigation for SARS-CoV-2 that did not have a documented RT-PCR result, which included asymptomatic patients who were exposed to someone with known COVID-19 and were monitored for symptoms, as well as patients who were diagnosed with COVID-19 outside the outpatient clinic.

We used the first reported suspicion date to define the day 0 among COVID-19 positive patients. In limited cases without a documented suspicion date, we used the RT-PCR positive test date to define the day 0. For a control group, we identified patients who had one or more negative COVID-19 test result without any positive result during the analysis period. We used the first COVID-19 negative test date to define the day 0 since the date of suspicion for negative patients was not recorded in the provider’s EHR. Patients with invalid/inconclusive test results were excluded from the analysis.

### Statistical Methods

We computed mean daily values for an array of clinical variables across the 90 days before day 0 for COVID-19 positive and COVID-19 negative groups. We reported findings from *a priori* selection of variables that appeared to have notable changes before COVID-19; these included pre-HD systolic blood pressure (SBP), pre-HD pulse, pre-HD body temperature, lymphocytes, neutrophil to lymphocyte ratio (NLR), ferritin, albumin, interdialytic weight gain (IDWG), and creatinine.

All data was collected during provision of standard medical care for HD patients. Data on SBP, pulse, body temperature, IDWG was collected on a per HD treatment basis. Laboratories were collected monthly with exception of ferritin that was collected on a quarterly basis.

Nonparametric smoothing splines^10^ were constructed to fit data for individual trajectories and estimate the mean change over time since first suspicion/COVID-19 positive or negative date. Among COVID-19 positive patients, we stratified data for those who survived or died within 30 days of first day 0; trajectories were plotted 90 days after day 0 in survivors and up to 30 days after day 0 in COVID-19 in patients who died.

For each clinical parameter of interest, the difference in average daily changes between COVID-19 positive versus negative group and COVID-19 survivor versus non-survivor group was estimated by fitting a linear mixed effects model based on measurements in the 14 days before (i.e., day-14 to day 0) day 0. The analysis used all available data without any imputation.

Average value of clinical and laboratory parameters on day 0 were compared using unpaired t-test. Average value of clinical and laboratory parameters on day 0 between survivors and non-survivors was also compared using unpaired t-test.

Analyses were performed using SAS version 9.4 (SAS, Cary, NC, USA). Smoothing splines and visualizations were conducted using R version 3.5.2 (R Foundation, Vienna, Austria).

## Results

### Characteristics of HD Patients who Received COVID-19 Testing

There were 12,836 HD patients with a suspicion of COVID-19 who received RT-PCR testing (8,895 COVID-19 positive and 3,941 COVID-19 negative patients) between 01 May and 01 Sep 2020. The demographics and comorbidities for the two groups of patients are shown in **Table 1**. There was a slightly lower proportion of patients with a white race and higher proportion of patients with diabetes in the COVID-19 positive group compared to the negative group. The mean number of days between the suspicion date and the positive test date in COVID-19 positive group was 5.6 days.

**Table 1:** Demographics and comorbidities of patients who tested COVID-19 positive and negative.

| Parameter | COVID-19 positive | COVID-19 negative |
| --- | --- | --- |
| Total number of patients | 8,895 | 3,941 |
| Male | 54% | 55% |
| White | 37% | 43% |
| Mean age as of first symptom date (std dev) | 61.8 (14.2) | 60.3 (14.7) |
| Mean vintage as of first symptom date (std dev) | 4.0 (4.1) | 3.8 (3.8) |
| Diabetes | 69% | 66% |
| Congestive Heart Failure | 21% | 22% |
| Ischemic Heart Disease | 24% | 24% |

### Trajectories of Vital Signs before COVID-19 Testing

We observed the COVID-19 positive group had decreases in pre-HD SBP weeks before day 0; the SBP was around 5 mmHg lower at the suspicion date versus 14 days prior (**Figure 1**). Contrary to this, the COVID-19 negative group had trends for increases in pre-HD SBP in the weeks before testing. The linear mixed effects model estimated the daily change in pre-HD SBP during the 14 days before day 0 and identified the COVID-19 positive group had an average decrease of −0.3 mmHg/day; this was distinct compared to the average increase of 0.2 mmHg/day found in the COVID-19 negative group (p<0.0001) (**Table 2**).

**Figure 1:**
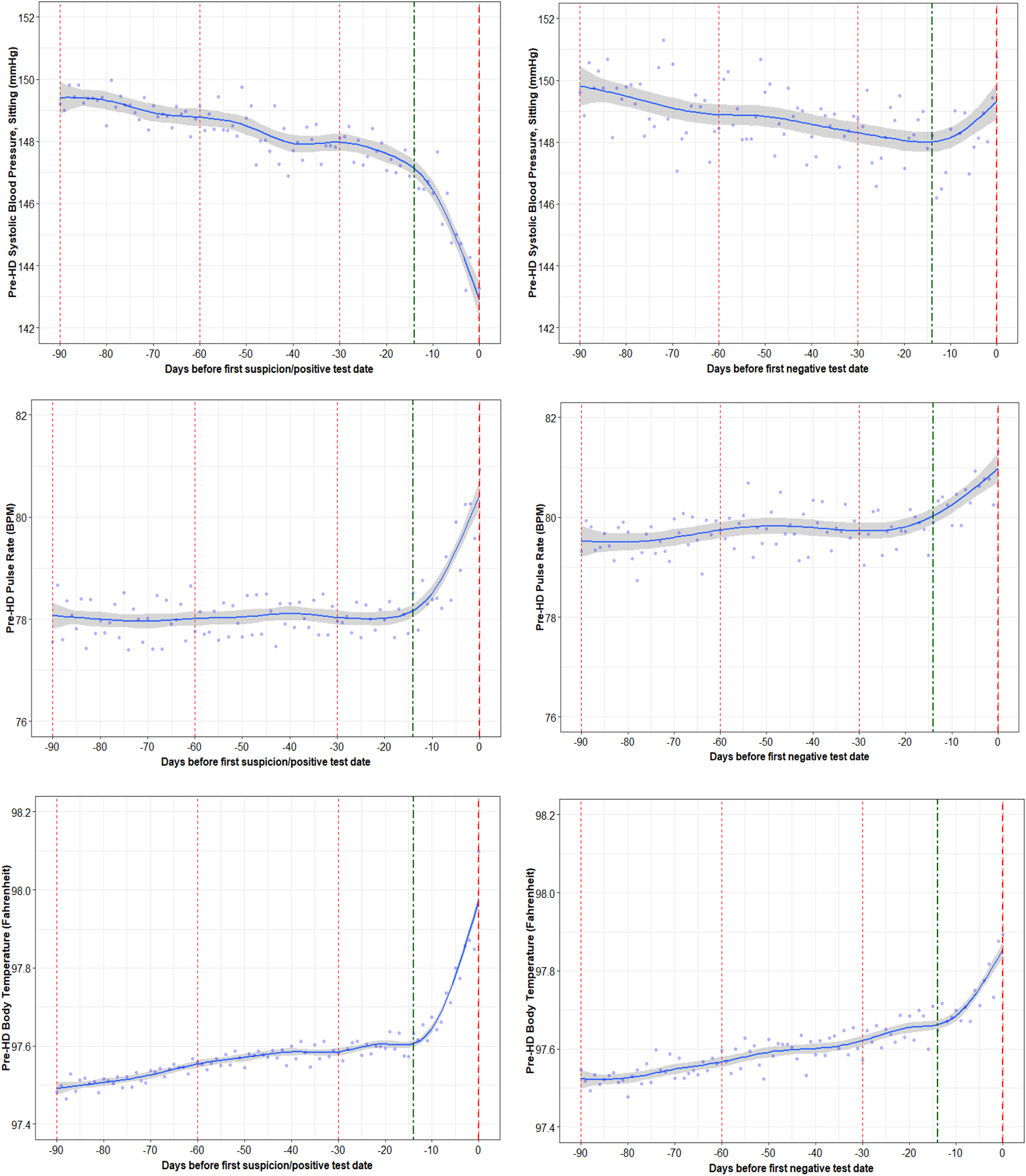
Trajectories of Pre-HD Vital Signs (SBP, Pulse Rate, Body Temperature) in COVID-19 Positive and Negative Patients

**Table 2:** Average daily change in clinical and laboratory parameters 14 days prior to day 0.

| Parameter | Mean Daily Change<br>COVID-19 positive | Mean Daily Change<br>COVID-19 negative | p-value |
| --- | --- | --- | --- |
| Pre-HD SBP (mmHg) | -0.2873 | 0.2293 | <0.0001 |
| Pre-HD Pulse Rate (BPM) | 0.1728 | 0.0880 | <0.0001 |
| Pre-HD Body Temp (F) | 0.0259 | 0.0140 | <0.0001 |
| Ferritin (ng/mL) | 26.1162 | 3.6907 | <0.0001 |
| Lymphocytes (%) | -0.1772 | -0.1208 | 0.0063 |
| Neutrophil-Lymphocyte Ratio(NLR) | 0.0907 | 0.0790 | 0.3359 |
| Albumin (g/dL) | -0.0123 | -0.0035 | 0.0004 |
| IDWG (kg) | -0.0441 | 0.0068 | <0.0001 |
| Creatinine (mg/dL) | 0.0127 | 0.0222 | 0.5561 |

*Average daily changes are compared using linear mixed effects model in 14 days prior day 0. \*P-value compares slopes in COVID-19 positive versus COVID-19 negative patients.*

The COVID-19 positive group was also found to have subtle increases in the pre-HD pulse and body temperature in the week before day 0. These trends were consistent with the COVID-19 negative group, but less pronounced. The average daily change in pulse rate and body temperature was found to be significantly larger in the COVID-19 positive versus COVID-19 negative group (p<0.0001).

The average values for the pre-HD SBP, pulse, and body temperature on day 0 is shown in **Table 3**. There were significant differences between groups for pre-HD SBP and body temperature (p<0.0001).

**Table 3:** Average value of clinical and laboratory parameters on day 0.

| **Parameter** | **COVID-19 positive** | **COVID-19 negative** | **p value** |
| --- | --- | --- | --- |
| **Pre-HD SBP (mmHg)** | 143.28 | 150.74 | <0.0001 |
| **Pre-HD Pulse Rate (BPM)** | 80.93 | 81.32 | 0.3266 |
| **Pre-HD Body Temp (F)** | 98.10 | 97.89 | <0.0001 |
| **Ferritin (ng/mL)** | 1500.78 | 1038.42 | 0.0004 |
| **Lymphocytes (%)** | 17.42 | 18.01 | 0.1794 |
| **Neutrophil-Lymphocyte Ratio (NLR)** | 5.53 | 5.40 | 0.6198 |
| **Albumin (g/dL)** | 3.52 | 3.63 | 0.0074 |
| **IDWG (kg)** | 1.53 | 2.30 | <0.0001 |
| **Creatinine (mg/dL)** | 9.04 | 8.76 | 0.3725 |

### Trajectories of Inflammatory Markers before COVID-19 Testing

Serum ferritin levels were found to have increased by around 400ng/ml in the COVID-19 positive group in the 14 days prior to day 0 (**Figure 2)**. There was a daily change in ferritin of 26.12ng/mL/day during the 14 days before day 0 in the COVID-19 positive group, which was distinct compared to the COVID-19 negative group that exhibited no remarkable changes (p<0.0001).

**Figure 2:**
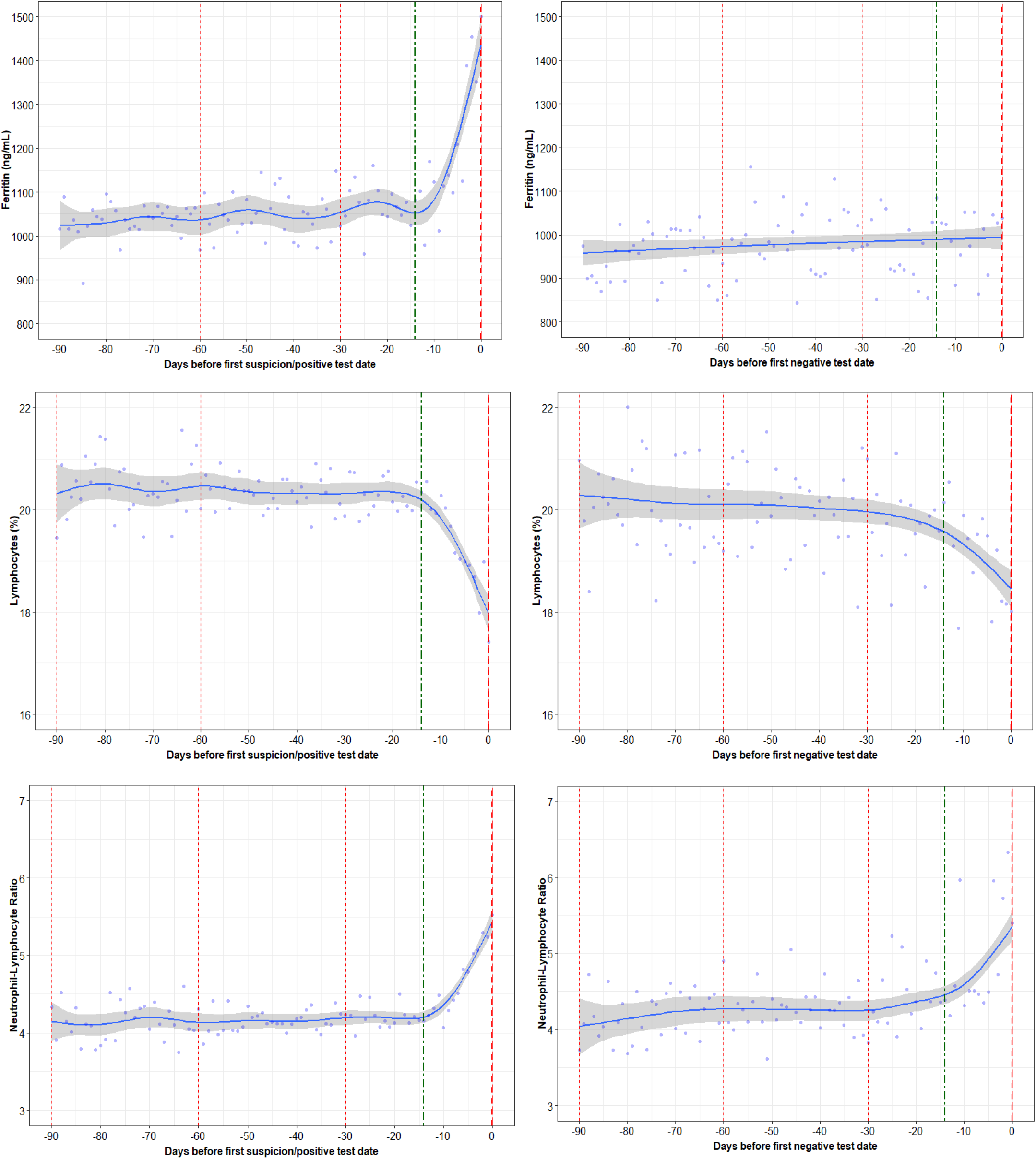
Trajectories in Inflammatory Markers (Ferritin, Lymphocytes, Neutrophil-Lymphocyte Ratio) in COVID-19 Positive and Negative Patients

The percentage of lymphocytes were found to decrease below 20% in both COVID-19 positive and negative groups with a significant difference in trends of daily change(p=0.0063). NLR showed an increasing trend in the COVID-19 positive and negative group 14 days prior to day 0, however, the difference was not significant.

On day 0, the only inflammatory marker that was different between groups was Ferritin, which was higher in the COVID-19 positive group (p=0.0004).

### Trajectories of Nutritional Markers before COVID-19 Testing

Among both groups, there was a decline in serum albumin in the 14 days prior to day 0, yet the decline was more pronounced in the COVID-19 positive group compared to COVID-19 negative group. The decline in albumin was an average of 0.012g/dL per day in the COVID-19 positive group compared to the decline of an average of 0.004 g/dL in the COVID-19 negative group (p=0.0004) **(Figure 3)**. Notably, IDWG decreased by of 0.6kg per day in the COVID-19 positive group in the 14 days prior to day 0 compared to almost no change in COVID-19 negative group (p<0.0001). There were no differences in the trends of creatinine between the COVID-19 positive and negative group.

**Figure 3:**
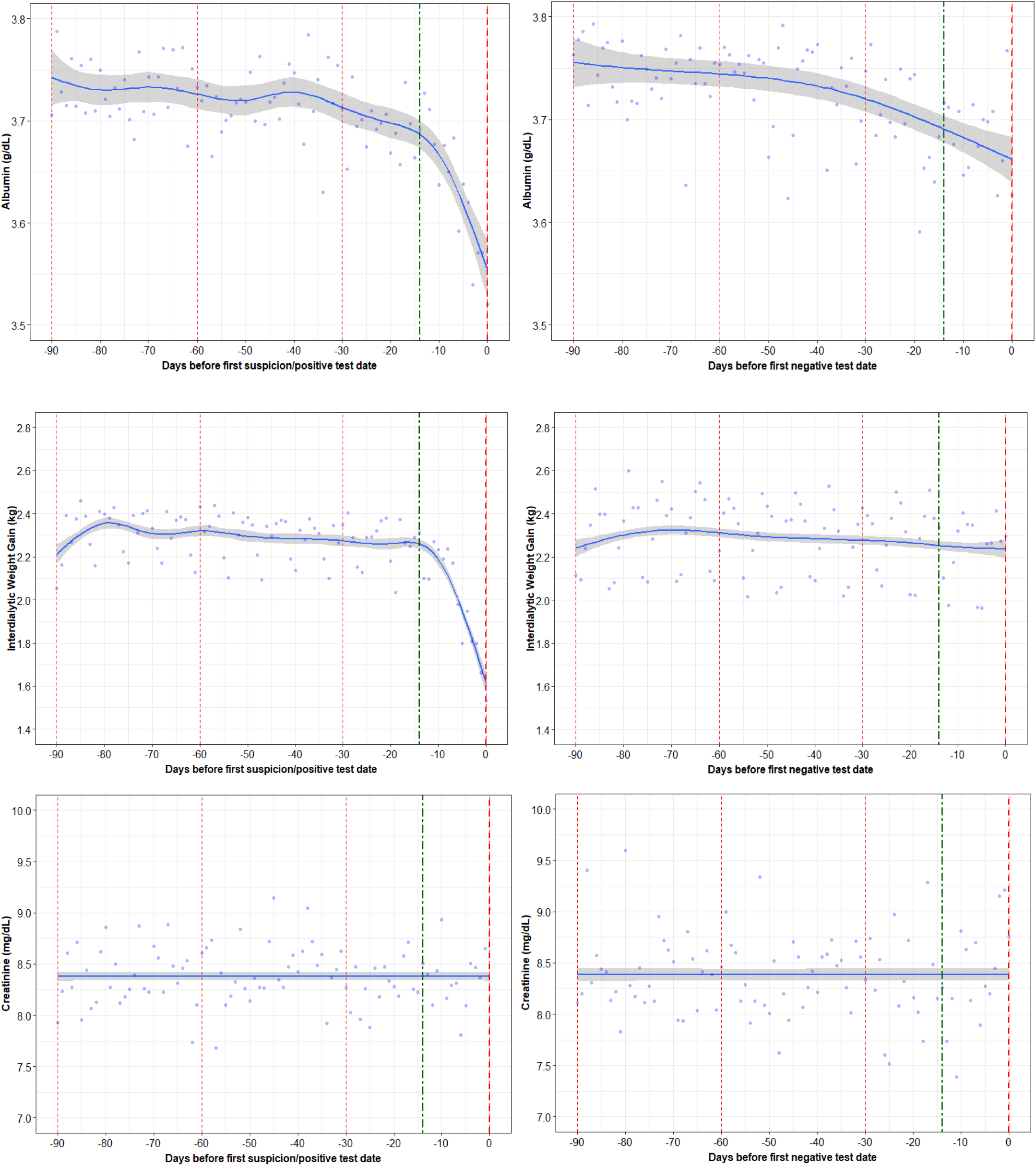
Trajectories in Nutritional Markers (Albumin, IDWG, Creatinine) in COVID-19 Positive and Negative Patients

Albumin and IDWG were significantly lower in the COVID-19 positive group on day 0 compared to COVID-19 negative group on the negative test date.

### Characteristics of HD Patients with COVID-19 by Survival

**Table 4** shows the demographics and comorbidities for patients diagnosed with COVID-19 who died within 30 days of COVID-19 positive date (non-survivors) versus those who survived (survivors). There were 7,897 survivors out of the 8,895 COVID-19 positive patients. The 998 non-survivors were more often older, male, white race, and had a higher comorbidity burden and longer dialysis vintage.

**Table 4:** Demographics and comorbidities of COVID-19 survivors and non-survivors.

| **Parameter** | **Survivors** | **Non-Survivors** |
| --- | --- | --- |
| **Total number of patients** | 7,897 | 998 |
| **Male** | 53% | 60% |
| **White** | 37% | 42% |
| **Mean age as of first symptom date (std dev)** | 60.8 (14.1) | 69.1 (12.5) |
| **Mean vintage as of first symptom date (std dev)** | 3.9 (4.1) | 4.7 (4.0) |
| **Diabetes** | 68% | 80% |
| **Congestive Heart Failure** | 21% | 28% |
| **Ischemic Heart Disease** | 23% | 29% |

### Trajectories of Vital Signs before and after COVID-19 by Survival

We observed a significant increase in the pre-HD pulse rate in the non-survivors versus the survivors with COVID-19. The linear mixed effects model showed an average increase of 0.29 BPM per day in the 14 days prior day 0 in the non-survivors compared to an average increase of 0.16BPM per day in survivors (p<0.0001) (**Table 5**). There was a difference in the pre-HD pulse rate between the survivors and non-survivors on day 0 (p=0.0528) (**Table 6**). Among survivors, it took 60 or more days after day 0 for pre-HD pulse to return to levels observed in the months before COVID-19.

**Table 5:** Average daily changes in clinical and laboratory parameters 14 days prior to day 0.

| Parameter | Survivors slope | Non-Survivors slope | P-value |
| --- | --- | --- | --- |
| Pre-HD SBP (mmHg) | -0.2786 | -0.3575 | 0.2222 |
| Pre-HD Pulse Rate (BPM) | 0.1582 | 0.2862 | <0.0001 |
| Pre-HD Body Temp (F) | 0.0261 | 0.0245 | 0.5362 |
| Ferritin (ng/mL) | 24.4168 | 41.1784 | 0.1998 |
| Lymphocytes (%) | -0.1569 | -0.3420 | <0.0001 |
| Neutrophil-Lymphocyte Ratio (NLR) | 0.0763 | 0.2067 | <0.0001 |
| Albumin (g/dL) | -0.0125 | -0.0126 | 0.9854 |
| IDWG (kg) | -0.0443 | -0.0438 | 0.9365 |
| Creatinine (mg/dL) | 0.0129 | 0.0067 | 0.8301 |

*Average daily changes are compared using linear mixed effects model in 14 days prior to day 0 in COVID-19 positive patients. \*P-value compares slopes in survivors versus non-survivors.*

**Table 6:** Average value of clinical and laboratory parameters on day 0.

| Parameter | Survivors | Non-Survivors | p value |
| --- | --- | --- | --- |
| Pre-HD SBP (mmHg) | 143.98 | 136.5 | 0.0002 |
| Pre-HD Pulse Rate (BPM) | 80.73 | 82.77 | 0.0528 |
| Pre-HD Body Temp (F) | 98.1 | 98.07 | 0.7363 |
| Ferritin (ng/mL) | 1497.37 | 1567.57 | 0.7386 |
| Lymphocytes (%) | 17.75 | 14.45 | 0.0038 |
| Neutrophil-Lymphocyte Ratio (NLR) | 5.3 | 7.53 | 0.0031 |
| Albumin (g/dL) | 3.54 | 3.21 | 0.0204 |
| IDWG (kg) | 1.56 | 1.26 | 0.0432 |
| Creatinine (mg/dL) | 9.06 | 8.68 | 0.7392 |

There difference in the daily change of pre-HD DBP and body temperature in survivors versus non-survivors during the 14 days prior to day 0 was not significant. On day 0 there were distinctions in pre-HD SBP between the survivors and non-survivors (p=0.0002). Among survivors, pre-HD SBP began to increase back up again around 10 days following day 0 and it took 30 days or more to return to levels observed before the infection; trends in non-survivors showed further and more pronounced decreases in pre-HD SBP during the 30 days after day 0 (**Figure 4**). There were no differences in pre-HD body temperature between the survivors and non-survivors on day 0 (p=0.7363). Pre-HD body temperature returned to levels seen in the months before COVID-19 around 60 days after day 0.

**Figure 4:**
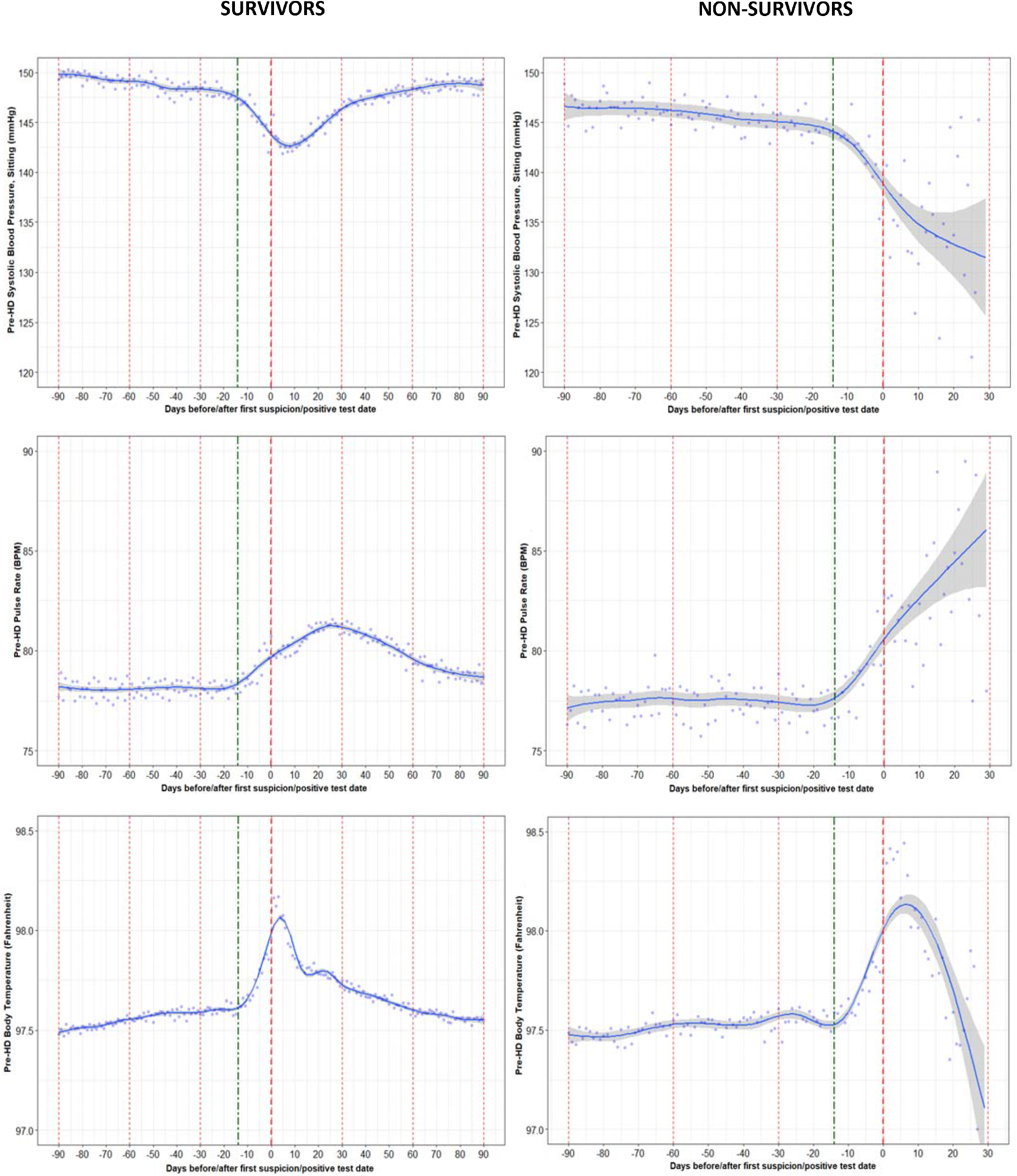
Trajectories in pre-HD Vital Signs (SBP, Pulse Rate, Body temperature) in COVID-19 Positive Survivors and Non-Survivors

### Trajectories of Inflammatory Markers before and after COVID-19 by Survival

There were significant differences in inflammatory markers like lymphocytes and NLR in both COVID-19 positive survivors and non-survivors. The average decrease in the percentage of lymphocyte was about 0.34% in the 14 days prior to day 0 in non-survivors compared to 0.16% in survivors (p<0.0001). Similarly, there was a difference in the 14-day trend for NLR between the survivors and non-survivors (p<0.0001). There were also significant differences in the percentage of lymphocytes and NLR between the survivors and non-survivors on day 0.

As shown in **Figure 5**, the disturbances in inflammatory markers returned to the levels seen in the months before the infection within about 30 days after day 0 in patients who survived. Patients who died experienced more robust decreasing trends in lymphocyte levels and thus a higher NLR in the 30 days following day 0 than those who survived.

**Figure 5:**
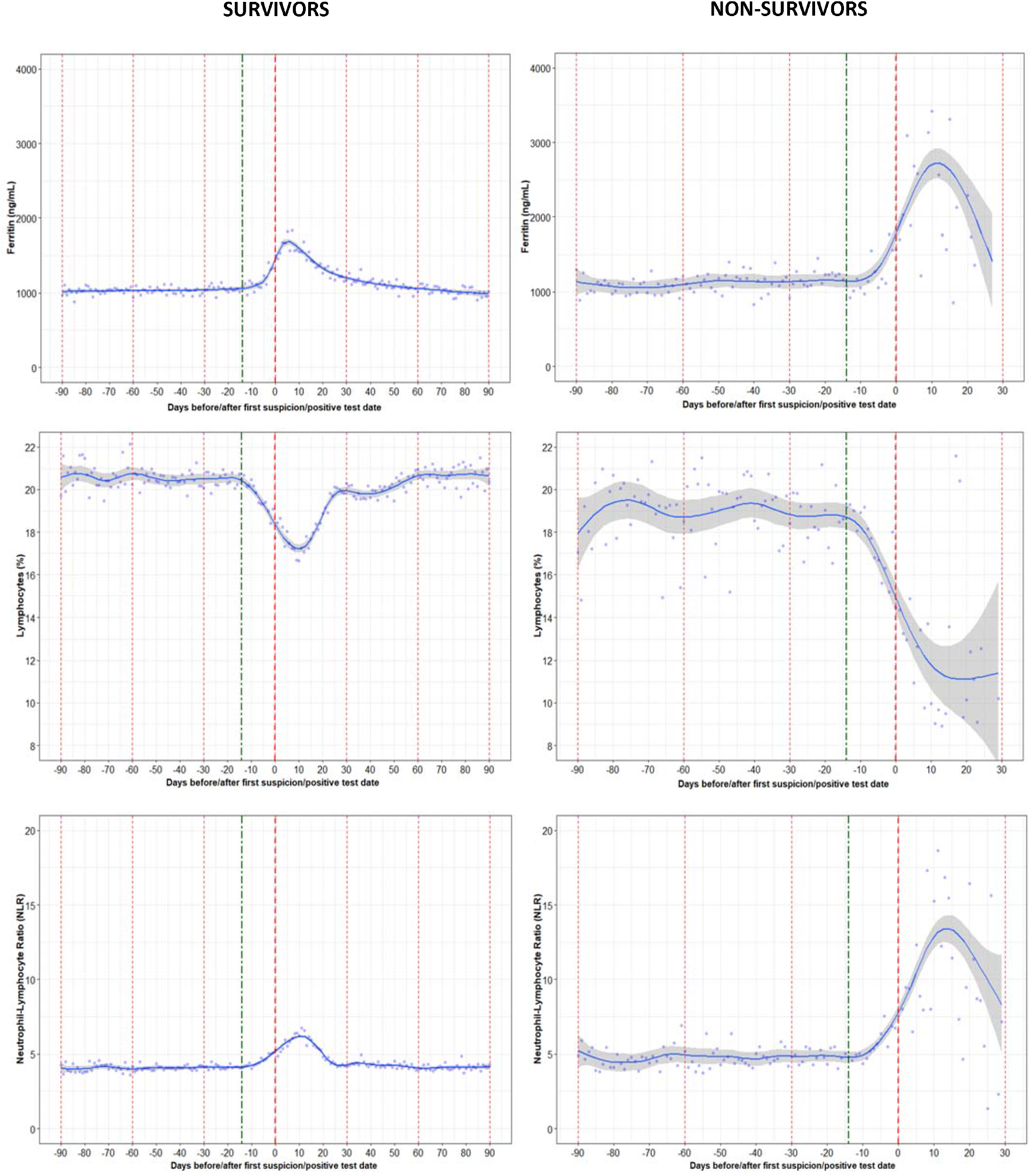
Trajectories in Inflammatory Markers (Ferritin, Lymphocytes, Neutrophil-Lymphocyte Ratio) in COVID-19 Positive Survivors and Non-Survivors

Difference in Ferritin between survivors and non-survivors was not significant during 14 days prior to day 0 and on day 0.

### Trajectories of Nutritional Markers before and after COVID-19 by Survival

Albumin levels among patients who died decreased steadily during the 30 days following day 0, whereas albumin levels began to rise after approximately 15 days following day 0 for patients who survived. Patients who died also experienced a larger decline in IDWG than the group who survived. Survivors experienced a decline in creatinine shortly after day 0, which remained lower than in the months before COVID-19 throughout the following 90 days. The average daily change was significantly different between groups for all nutritional markers (albumin and IDWG) in the 14 day prior to day 0 test date between the survivors and non-survivors (p<0.05). The difference in the slope of creatinine between the survivors and non-survivors was not significant(p=0.7392).

There were also significant differences in albumin and IDWG between the survivors and non-survivors on day 0. Among survivors, the nutritional parameters took around 60-90 days to return to the same levels as in the months before the infection (**Figure 6**).

**Figure 6:**
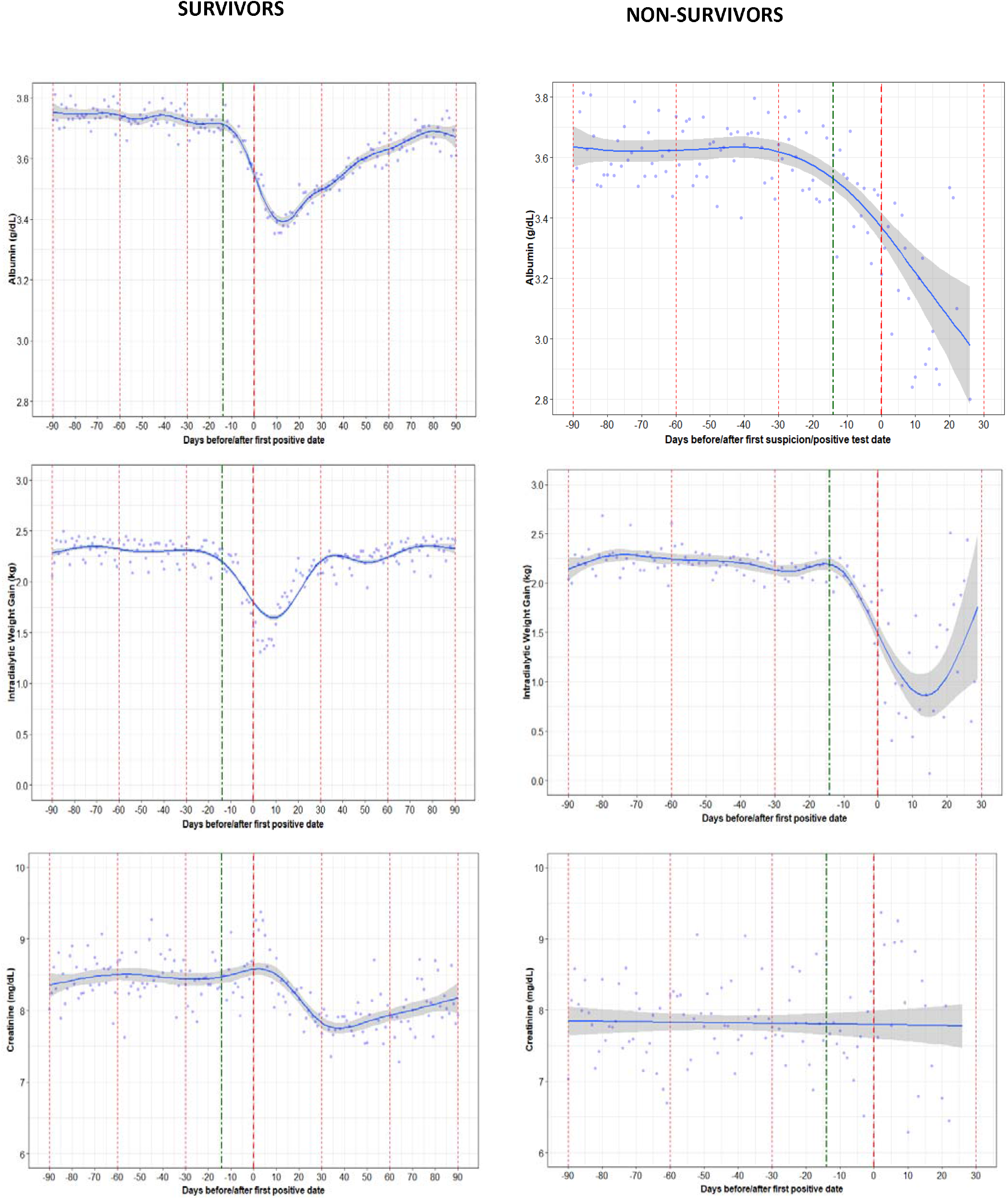
Trajectories in Nutritional Markers (Albumin, IDWG, Creatinine) in COVID-19 Positive Survivors and Non-Survivors

## Discussion

We observed unique temporal trends in various clinical and laboratory parameters among HD patients who tested positive versus negative for SARS-CoV-2 infection. Ultimately, these trends help to define the physiological disturbances that characterize the onset and course of COVID-19 in HD patients. The disturbances in various parameters commonly started around 14 days before presentation and were more pronounced in patients who tested positive for COVID-19. We identified statistically distinct daily changes in vital signs, inflammatory, and nutritional markers across the weeks before presentation among patients who tested COVID-19 positive versus negative, as well as unique differences between groups on the date of suspicion/testing. Albeit statistically significant, the differences were often clinically subtle for many parameters.

Nonetheless, these findings are anticipated to be of use in development of diagnostic support risk scores and prediction models. Among patients in the COVID-19 negative group, there were observed in various parameters which were anticipated since these patients likely underwent testing due to presentation with symptoms of a flu-like illness; however, we cannot exclude that some were tested secondary to exposure to someone with known COVID-19. Among the COVID-19 positive group, we identified clinically remarkable disturbances in vital signs, inflammatory, and nutritional markers after being diagnosed with COVID-19 that commonly took more than one month to return to normal among those who survived. The trajectories in parameters were distinct between survivors and non-survivors brining insights that will be of importance to consider in the development of prognostic support risk scores and prediction models.

Published studies showing clinical trends before and at onset in COVID-19 HD patients are scarce. We recently showed how a machine learning predictive model that uses changes in clinical parameters before diagnosis had reasonable performance in classification of patients with a SARS-CoV-2 infection three days before symptoms onset (area under the curve is the testing dataset was 0.68). The model developed showed changes in IDWG had the highest variable feature importance (reflecting the impact of the value to the prediction) ^11^. Our present analysis adds to previous findings by showing quantitative trends in key parameters during the period before and after presentation with COVID-19. One of the novel findings we identified was the inverse trends in pre-HD SBP before presentation between COVID-19 positive versus negative patients. The decreases in SBP specifically associated with the onset of COVID-19 may be representative of some potential direct or indirect influences of the disease on the heart that caused a decompensation in cardiovascular system. The specificity of trends in physiological parameters for COVID-19 diagnosis should be assessed in future studies.

When comparing survivors and non-survivors, we found that mean age, dialysis vintage, and the proportion of males was higher in non-survivors, as was the presence of comorbidities such as diabetes and congestive heart failure and this consistent with previous findings ^3, 4, 12^. Previous studies also have showed that higher body temperature, lower lymphocyte counts, higher levels of C-reactive protein and white blood cell counts were related to mortality in HD patients ^3, 4, 12-15^. However, we observed that average change in temperature 14 days prior to day 0 was not significantly different in survivors compared to non-survivors. The body temperature on day 0 for non-survivors was also slightly higher than survivors, although the difference was not significant.

In patients testing positive for COVID-19, the laboratory parameters anticipated to be directly related to the infection, such as NLR, lymphocytes, and ferritin, returned to their baseline values within 30 days after suspicion/testing. The relatively long time for infection related parameters to persist may be because SARS-CoV-2 infection may persist for longer period in HD patients. In a study of 19 HD patients with COVID-19 with repeat RT-PCR testing, SARS-CoV-2 tests remained positive in 68% patients after 20 days and in 32% after 40 days ^16^.

We found a modest decline in IDWG in the weeks before presentation that was specific to the COVID-19 positive group. Among patients who contracted COVID-19, the decline in IDWG was more pronounced in non-survivors versus survivors, which may be a prognostic signal for malnutrition when accompanied by a deterioration in other nutritional parameters ^17^. Parameters related to nutrition appear to take longer to return to the baseline in patients who survived suggesting the systemic effects of COVID-19 may be prolonged. Serum creatinine, which can also be considered a marker of lean tissue mass ^18^, even had not return to baseline 90 days after diagnosis. This suggests profound catabolic effects of COVID-19, which are due to multiple mechanisms such as anorexia, hypoxia, immobilization and increased levels of pro-inflammatory cytokines ^19^. In our analysis, it also took 2-3 months for serum albumin to return to the baseline values seen in the months before infection.

This analysis assessed a large group of COVID-19 positive HD patients and provides novel information. However, it is important to note that patients could have been tested for presentation with symptoms or exposure to someone with known COVID-19, which cannot be fully deduced in cases where there is missing/unavailable data on the date of suspicion. Although most patients were likely tested for clinical reasons, which is known for most of the COVID-19 positive patients, this is a limitation of the analysis. In general, absolute differences in trends, although significant, were small and we therefore suggest using multiple markers in combination for risk prediction, as shown in our previous paper regarding a machine learning prediction model developed for early detection of patients with COVID-19 ^11^. Moreover, it is important to realize that the trends do not show the mean of individual trajectories, but an aggregate of the cohort groups. We cannot rule out that there might be some minimal temporal bias secondary to the definition of the reference date/day 0 for suspicion/testing that may have an impact on the trajectories. Also, there is a possibility that the incubation period of the virus was prolonged due to low immunity in dialysis patients. Nonetheless, the comparisons in trends in daily changes during the weeks prior suspicion/testing are anticipated to have reasonably captured signals. Further investigations should consider inclusion of these insights to improve the precision of COVID-19 risk scores and prediction models being developed and used in care paradigms.

## Conclusion

We found the trajectories of several clinical/laboratory parameters distinctly change in the weeks before presentation with COVID-19, as compared to patients who were tested for a SARS-CoV-2 infection and found negative. Among patients who were diagnosed with COVID-19, the survivors appeared to have distinct trajectories in clinical/laboratory parameters compared to patients who died within 30 days after COVID-19 suspicion/testing. These findings appear to reveal some of the pathophysiologic trends defining the onset and course of the disease in the HD population, however, many changes were small. In patients with COVID-19 who survived, inflammatory and cardiovascular parameters returned to baseline levels in 1 month, however, the effect of COVID-19 on nutritional status appears to be more prolonged. These insights are anticipated to be of high importance for development of prediction models for early identification and prognosis of COVID-19.

## Data Availability

This analysis used data from patients treated at an integrated kidney disease company in the United States (Fresenius Medical Care North America). The data was deidentified by the investigator and is restricted to use by authorized individuals. The deidentified dataset is not publicly available.

## Disclosures

SC is a student at Maastricht University Medical Center. SC, RL, YJ, JL, CM, AW, LN, JH, LU and FM are employees of Fresenius Medical Care. PK is an employee of Renal Research Institute, a wholly owned subsidiary of Fresenius Medical Care. SC, PK, JH, LU, FM have share options/ownership in Fresenius Medical Care. PK is an inventor on multiple patents in the field of dialysis and is on the Editorial Board of Blood Purification and Kidney and Blood Pressure Research. FM has directorships in the Fresenius Medical Care Management Board, Goldfinch Bio, and Vifor Fresenius Medical Care Renal Pharma. SL, YW and JPK has no relevant conflicts of interest to disclose.

## Funding

Analysis and manuscript composition were supported by Fresenius Medical Care.

## Authors’ contributions

Design was performed by SC, RL, YJ, JL, CM, AW, LN, PK, SL, YW, JPK and LU. Data extraction and analysis was performed by SC, RL, YJ, and LU. The interpretation, drafting and revision of this manuscript was conducted by all authors. The decision to submit this manuscript for publication was jointly made by all authors and the manuscript was confirmed to be accurate and approved by all authors.

